# REAL TIME MONITORING OF RESPIRATORY VIRAL INFECTIONS IN COHORT STUDIES USING A SMARTPHONE APP

**DOI:** 10.1101/2024.04.03.24304240

**Authors:** David G Hancock, Elizabeth Kicic-Starcevich, Thijs Sondag, Rael Rivers, Kate McGee, Yuliya V Karpievitch, Nina D’Vaz, Patricia Agudelo-Romero, Jose A Caparros-Martin, Thomas Iosifidis, Anthony Kicic, Stephen M Stick

**Affiliations:** Wal-yan Respiratory Research Centre, Telethon Kids Institute, Nedlands, 6009, Western Australia, Australia; Department of Respiratory and Sleep Medicine, Perth Children’s Hospital, Nedlands, 6009, Western Australia, Australia; WeGuide, The Royal Children’s Hospital, Parkville 3052, Victoria, Australia; School of Population Health, Curtin University, Bentley, 6102, Western Australia, Australia; Centre for Cell Therapy and Regenerative Medicine, School of Medicine, The University of Western Australia, Nedlands, 6009, Western Australia, Australia; European Virus Bioinformatics Centre, Jena, Thuringia, Germany; School of Biomedical Sciences, The University of Western Australia, Nedlands, 6009, Western Australia, Australia

**Keywords:** Paediatric Lung Disease, respiratory infections (non-tuberculous), viral infection

## Abstract

**Background and Objectives:** Cohort studies investigating respiratory disease pathogenesis aim to pair mechanistic investigations with longitudinal virus detection but are limited by the burden of methods tracking illness over time. In this study, we explored the utility of a smartphone app to robustly identify symptomatic respiratory illnesses, while reducing burden and facilitating real-time data collection and adherence monitoring.

**Methods:** The AERIAL TempTracker smartphone app was assessed in the AERIAL and COCOON birth cohort studies. Participants recorded daily temperatures and associated symptoms/medications in TempTracker for 6-months, with daily use adherence measured over this period. Regular participant feedback was collected at quarterly study visits. Symptomatic respiratory illnesses meeting study criteria prompted an automated app alert and collection of a nose/throat swab for testing of eight respiratory viruses.

**Results:** In total, 32,764 daily TempTracker entries from 348 AERIAL participants and 30,542 entries from 361 COCOON participants were recorded. This corresponded to an adherence median of 67.0% (range 1.9-100%) and 55.4% (range 1.1-100%) of each participant’s study period, respectively. Feedback was positive, with 75.5% of responding families reporting no barriers to use. A total of 648 symptomatic respiratory illness events from 249/709 participants were identified with significant variability between individuals in the frequency (0-16 events per participant), duration (1-13 days), and virus detected (rhinovirus in 42.7%).

**Conclusions:** A smartphone app provides a reliable method to capture the longitudinal virus data in cohort studies which facilitates the understanding of early life infections in chronic respiratory disease development.

**Summary at a Glance:** A smartphone app can facilitate capturing symptomatic respiratory viral infections in longitudinal cohort studies, while supporting adherence and reducing participant burden. The app helped identify community variations in virus prevalence as well as the individual variability in viral responses necessary to understand the mechanism of chronic disease development.

## INTRODUCTION

The pathogenesis of human respiratory disease represents a complex interplay between intrinsic and extrinsic factors, with respiratory virus exposures playing a critical role in their development and manifestations^1–4^. While some individuals experience recurrent, severe viral infections and develop chronic lung disease, other individuals rarely develop symptomatic illness, even when infected with the same virus^1–4^. Understanding the biological mechanisms driving this variability remains a key research priority^5^. To address this, multiple cohort studies have aimed to pair mechanistic investigations with longitudinal tracking of individual health trajectories, with different approaches taken to characterise infection patterns over time^6–12^.

Significant respiratory illness events can be identified by monitoring hospital and/or primary healthcare attendance^13^, but this approach works best in regions with centralised healthcare and will miss relevant illnesses not requiring medical attention. Questionnaires and interviews can retrospectively capture illness events but are subject to recall bias particularly when performed infrequently^14,15^. Another approach is to use regular study visits and/or baseline virus sampling to capture illnesses^12^, with more frequent interactions decreasing the likelihood of missing events but increasing resource utilisation and participant burden. Finally, symptom diaries have been frequently utilised for either retrospective identification of illness episodes or as a tool to promote reporting of events^8^. However, traditional paper-based symptom diaries are often onerous on participants and/or can be inconsistently completed^14,16–18^. Novel approaches to enable robust illness detection with reduced participant and resource burden are still needed.

The utilisation of smartphone-based technologies has several benefits including improved usability, integration into existing use patterns, built-in app alerts, and easier facilitation of remote data management and adherence monitoring^10,19–22^. While smartphone apps have been increasingly used in research^10,19–22^, their application for symptomatic respiratory illness detection in cohort studies requires further exploration. Here, we present the development, implementation, and validation of an in-house smartphone app named AERIAL TempTracker and its real-world application in two cohort studies^5^.

## METHODS

### Consumer Consultation and Co-Design

To ensure high integrity of our cohort studies, we consulted two consumer reference groups representing Telethon Kids Institute Respiratory Research and ORIGINS Project. All consumers insisted on simplicity and ease of monitoring, communication, and daily recordings to reduce burden. Consumers also suggested avoiding email/text message reminders and pen and paper documentation, which led to the development of the AERIAL TempTracker smartphone app.

Once the app build was complete, the same consumers and the wider community were once again invited to test and evaluate the prototype app to provide input and feedback on the following aspects of the app: first impressions, design, content, functionality and flow, features, technical issues, likes and dislikes. The feedback was collated and discussed with the design team and the changes incorporated into the app by the developers (Supplementary Table 1).

### TempTracker development and workflow

TempTracker, available on iOS and Android, was developed by WeGuide Pty Ltd, a subsidiary of Curve Tomorrow Pty Ltd. To adhere to ethics and governance procedures, TempTracker integrated with a Research Electronic Data Capture (REDCap) database^23,24^ located on secured Telethon Kids Institute servers, providing real-time data monitoring. Participant families were able to use the app on multiple devices with the same login, with all recruited participants in the same family able to be added to the same device or multiple devices, depending on preference.

TempTracker’s workflow is presented in Figure 1 and Supplementary Figure 1. For each entry, participants record (1) measurement time; (2) temperature; (3) symptoms present; (4) medications used; and optionally (5) other free-text symptoms/medications. Recorded temperatures over 37.5°C were prompted to be re-taken. Events meeting pre-programed criteria (detailed below) triggered an automated alert to the participant and study team for respiratory virus testing. TempTracker provided a daily reminder, with the notification time set by the family.

**Figure 1.**
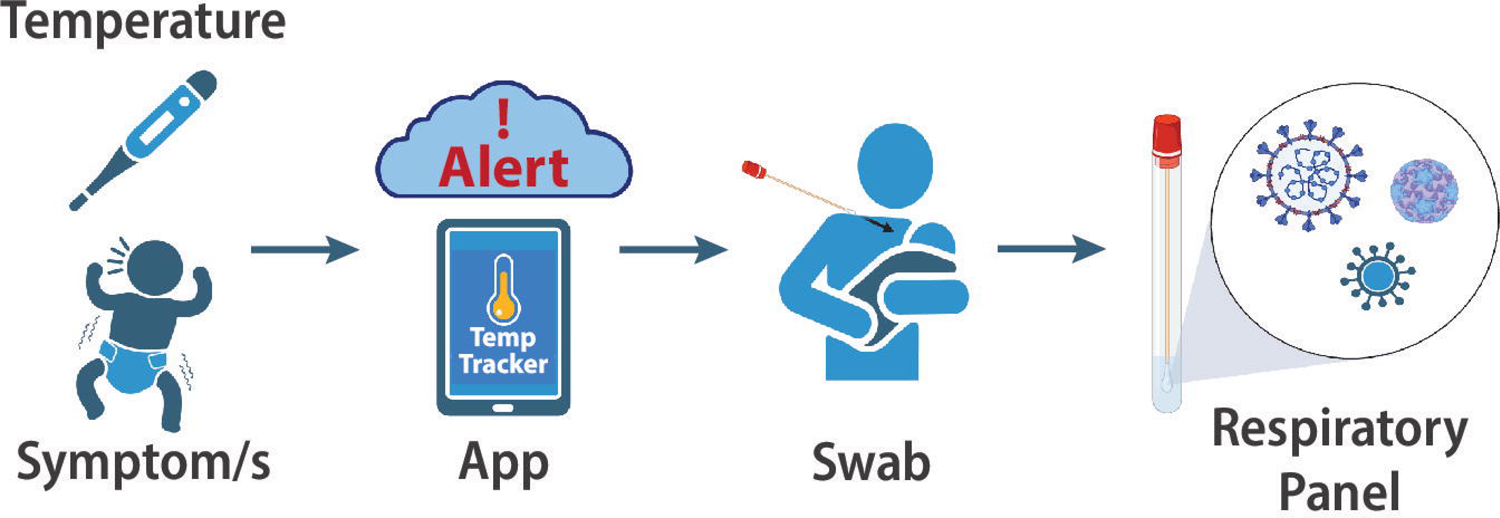
AERIAL TempTracker smartphone app schema for the AERIAL and COCOON cohort studies. Participants (or adult family members of consented participants) entered daily temperatures and symptoms into the AERIAL TempTracker smartphone app. Events meeting the criteria for a symptomatic respiratory illness generated an automated app alert to perform a nose/throat swab for respiratory virus detection.

### Study Settings

TempTracker was used in two sub-studies nested within the ORIGINS Project, a large longitudinal, prospective birth cohort study based at Joondalup Health Campus, Western Australia^25,26^. AERIAL (<u>A</u>irway <u>E</u>pithelium <u>R</u>espiratory Illnesses and <u>AL</u>lergy Study) recruited mother/newborn dyads to determine the influence of early-life viral infections on asthma/atopy development^5^. The study population consisted of consented newborn participants whose parent(s) entered their data into TempTracker. COCOON (<u>CO</u>VID <u>CO</u>mmunity compassi<u>ON</u> Study) recruited complete family units to elucidate patterns of household transmission during the COVID-19 pandemic. The study population consisted of all consented family members (newborns, parents, other children, and other adults), with one or more family members (usually parents) entering their family’s data. Recruitment commenced in August 2020 during the COVID-19 pandemic and the cohorts were required to adhere to evolving Western Australian Health Department guidelines.

### Symptomatic Respiratory Illness Detection

Families were asked to measure daily temperatures for 6-months using the provided digital forehead thermometer and record the temperature and associated symptoms/medications in the TempTracker app for each enrolled participant (newborn in AERIAL, all family members in COCOON). After the initial 6-months, participants continued with weekly temperature measurements and app use (data not analysed here) “Symptomatic respiratory illness” events meeting the study criteria were identified in TempTracker with pre-programmed logic and defined as a temperature >37.5°C and the presence of at least one respiratory symptom, or temperature <37.5°C and the presence of at least two respiratory symptoms (Supplementary Table 2). These criteria were developed in accordance with contemporaneous Western Australian health guidelines for symptomatic severe acute respiratory syndrome-related coronavirus-2 (SARS-CoV-2) testing.

Participants meeting the study criteria had their throat and one nostril^27^ sampled with a swab (FLoQSwabs, Copan Group, Italy), which was placed into viral transport medium (eSwab, Copan Group, Italy) and stored at 4°C until transport to a commercial laboratory (Westerns Diagnostics Pathology, Jandakot, Western Australia, Australia) for testing against a panel of eight respiratory viruses (influenza ‘A’, influenza ‘B’, respiratory syncytial virus (RSV), human metapneumovirus, parainfluenza, rhinovirus, adenovirus, SARS-CoV-2) by multiplex polymerase chain reaction. Symptomatic swabs were only taken if the participant/parent gave consent and if their previous swab was more than two-weeks prior. Swabs were initially done by study staff at home visits; however, the protocol was adjusted in March 2022 to enable parent collection to adhere to public health restrictions. All virus results were shared with participant families.

Influenza A and RSV detections in AERIAL and COCOON cohorts were compared to wider Western Australian community viral transmission data extracted from the Western Australian Department of Health’s paediatric respiratory pathogen reports^28^.

### Usage Analysis

Participants were required to login to TempTracker to qualify as “activated” users. For each participant, their eligible study period was defined as the time between app activation and either six months or the date the participant was lost to follow-up or withdrew. While participants were requested to use TempTracker daily, they were still considered “active” users if they used TempTracker at least once every 7 days, given the lower chance of missing respiratory events. Participant feedback on TempTracker was captured as part of a routine quarterly questionnaire for both studies.

## RESULTS

### AERIAL Study App Usage

Between September 2020 and April 2023, the AERIAL birth cohort study recruited 361 newborn participants from 358 families. Thirteen participants (3.6%) were never activated on TempTracker yet remained adherent with other study procedures. Of the remaining 348 participants (96.4%), 304 (87.4%) continued in the study throughout the 6-month study period, while 44 (12.6%) were either lost to follow-up (n=36) or formally withdrew (n=8) at a median age of 2.7 months (range 0.6 – 4.9 months).

A total of 32,764 daily entries were recorded. TempTracker was used on a median of 67.0% (range 1.9-100%) of each participant’s eligible study days (Figure2A). Over the 6-month analysis period, 229 newborn participants (66.0%) remained “active” users (i.e. recording entries at least once every 7 days) for more than 80% of their study period (Figure2B).

### COCOON Study App Usage

Between August 2020 and October 2021, the COCOON cohort study recruited 101 participant families comprising a total of 419 individuals, including 99 (23.6%) newborns, 78 (18.6%) other siblings/children, 101 (24.1%) mothers, 94 (22.4%) fathers/partners, and 47 (11.2%) other adults (i.e. grandparents, great-grandparents, uncles). The median family size was 4 persons (range 3 to 8). In total, 58 (13.8%) recruited individuals (7 newborns, 3 other children, 4 mothers, 6 fathers/partners, and 38 other adults) were never activated on TempTracker. Of the 361 (86.2%) participants that were activated on TempTracker, 336 (92.8%) continued in the study through the 6-month study period, while 26 (7.2%) participants were either lost to follow-up (19 participants from 5 families) or formally withdrew (7 participants from 2 families) at a median of 2.2 months (range 0.5 – 5.0 months).

A total of 30,542 daily entries were recorded. TempTracker was used with a median of 55.4% (range 1.1-100%) of each participant’s eligible study days (Figure2C). A higher frequency of daily use was observed for mother (61.1%) and newborn (59.7%) participants whereas other family members used TempTracker less frequently (fathers/partners (39.2%), other children (44.4%), and other adults (18.3%)). Over the 6-month analysis period, 195 participants (54.1%) remained “active” users for more than 80% of their study period (Figure2D).

### Participant Feedback

Of the 451 families from both AERIAL and COCOON, 111 (24.6%) provided a response on the benefits of TempTracker use with receiving viral testing results (n=85, 76.6%) being the most frequently reported. More feedback was received for barriers to regular use with 326 (72.2%) families provided 496 total responses. Most families (n=246, 75.5%) did not report any issues with daily temperature and symptom recording in TempTracker. However, at least one barrier was reported by 80 (24.5%) participant families, and included burden of daily temperature measurements (n=33), general forgetfulness (n=21), maternal return to work (n=14), logistics of completing study requirements (e.g. family/holidays/moving; n=10) and issues with their smartphone (n=7), thermometer (n=7), or TempTracker (n=4). Maternal return to work was primarily reported at the end of the 6-month study period (n=12), with only 2 participant families reporting this reason earlier at 3.3 and 4.5 months of age, respectively.

Participant families were also asked whether they were willing to use TempTracker beyond the initial study period, with 324 out of 451 families (71.8%) providing a response. Of those responding, 223 (68.8%) were happy to continue with daily or second daily use, 46 (14.2%) preferred weekly use, and 27 (8.3%) preferred use only on days when unwell or symptomatic, while 28 (8.6%) did not want to continue.

### Symptomatic Respiratory Illnesses

Across the combined 63,306 daily entries from both studies, the mean recorded temperature was 36.7°C (range 35.0-39.9°C). Elevated temperatures (>37.5°C) were recorded in 878 (1.4%) app usages, with 130 (0.2%) above 38°C.

A total of 1,523 distinct symptomatic events (consecutive days with any positive symptom or temperature >37.5°C) were detected from 428/707 (60.4%) participants (median of 2 per participant, range 1-24). These events covered 3,583 total symptomatic days, with each event lasting a median of 1 day (range 1-67 days). Although TempTracker’s free text field was only used in 235 total app entries (0.37%), it captured additional health data such as positive SARS-CoV-2 rapid antigen tests (n=41, 17.4%), possible vaccine reactions (n=32, 13.6%), urinary tract infections (n=14, 5.9%), and meningitis diagnoses (n=3, 1.3%).

Of the 1,523 symptomatic events, 648 (42.5%) events from 249 participants (median 2 per participant, range 1-16) met our study criteria as “symptomatic respiratory illnesses” (Methods). These covered a total of 1,304 illness days with a median duration of 1 day (range 1-13 days). The most frequently reported respiratory symptoms were blocked nose (n=448, 69.1%), sneezing (n=301, 46.5%), runny nose (n=287, 44.3%), dry cough (n=268, 41.4%), and wet cough (n=167, 25.8%). Medication use was recorded in 410 (63.3%) events, with antipyretics (ibuprofen/paracetamol; n=345, 53.2%) and decongestants (n=89, 13.7%) being the most common.

Over the study period, 374 symptomatic swabs were performed on 234 participants, with respiratory viruses detected on 254 (67.9%) of the swabs. Rhinovirus was the most common virus detected (n=167, 42.7%), followed by RSV (n=33, 8.8%), SARS-CoV2 (n=32, 8.6%), human metapneumovirus (n=18, 4.8%), parainfluenza (n=17, 4.5%), influenza A (n=7, 1.9%), and adenovirus (n=2, 0.5%). No cases of influenza B were detected. The prevalence of influenza A (Figure3A) and RSV(Figure3B) within AERIAL and COCOON matched those reported in public health data for the same period.

In COCOON, 26 household transmission events were identified with multiple (range 2-5) family members concurrently meeting the study criteria. At least one virus was detected in 22 (84.6%) of these events, with rhinovirus (n=16, 61.5%) and RSV (n=4, 15.4%) the most prevalent. In all transmission events there was significant variability in symptom burden between participants, and in in all but one event (n=25, 96.2%) there was at least one additional family member (range 1-4) that did not become either symptomatic or virus positive.

## DISCUSSION

Accurate respiratory viral illness detection is essential for understanding the pathogenesis and progression of respiratory disease in longitudinal cohort studies. However, conventional methods are often labour intensive, placing a significant burden on participants, study staff, and resources. In this study, we have shown that a smartphone app can facilitate longitudinal symptomatic virus detection, support adherence, and provide meaningful data across two cohort studies.

Reducing participant burden has been well documented as a critical factor in maximising cohort study retention and robustness of data^29^. Consistent with this, the consulted consumer reference groups highlighted usability and decreased burden as their key priority areas, with the AERIAL TempTracker smartphone app being the key outcome of this co-design. In both cohorts, we had strong uptake of TempTracker as well as excellent retention through 6-months. We also observed a strong commitment for daily recording, with 66.0% of AERIAL participants and 54.1% of COCOON participants remaining “active” users. This aligns with previous studies tracking respiratory symptoms or viruses over time which report highly variable adherence ranging from 10-90%, with duration of follow-up, frequency of contact, and study population being key factors^8,12,18^.

One integral benefit of TempTracker was its ability to generate daily usage reminders and track adherence in real-time, with previous studies relying on either regular participant contact and/or retrospective review to monitor adherence^6–9,18^. Automated alerts through text-messages or emails have also been increasingly used to prompt study activities^17,30^, but a dedicated smartphone app is necessary to pair reminders with data collection/monitoring. Another major benefit of a smartphone app is the ability to pre-program logic to define an event and automate “take-a-sample” alerts, rather than relying on participant education and recall for sampling^14,16,17^. This enables standardised study criteria, while reducing missed sampling of true events or unnecessary additional sampling, particularly in cases where health literacy may be limited^31^. With increasing smartphone usage in society^32^, another advantage is the natural integration into regular usage patterns. However, it is important to note that smartphone-based technologies are not suitable for all potential users, and so mixed online/offline resources may be required^8^. This was observed in AERIAL, where one family did not own a device capable of supporting TempTracker.

Overall feedback was positive with most families reporting no issues and a willingness to continue with TempTracker. Obtaining viral testing results when unwell was highlighted as the primary benefit and likely contributed to the strong adherence by empowering families with viral results to counteract the well-documented feelings of uncertainty, fear, and anxiety around respiratory infection during the pandemic^33–36^. Barriers to TempTracker usage were reported by a quarter of families, with the most frequent being the burden of daily temperature measurements. Alternate approaches such as measuring temperatures only when symptomatic or removing temperatures entirely may have increased adherence, particularly as temperatures >38°C were only recorded in 0.2% of all entries. However, we identified multiple instances of positive viral detection in the context of low-grade fever but minimal symptoms, supporting the rationale for their inclusion. Maternal return to work was reported as another common barrier, primarily at 6-months when more mothers had returned to the workforce. This barrier was not unexpected as return to work has been well-described as a critical timepoint with wide-reaching impacts on both maternal and infant health outcomes, as well as influencing factors such as immunisation uptake and breastfeeding duration^37^.

Across the symptomatic respiratory illnesses meeting our study criteria, we observed significant variation in the number of illnesses each participant experienced, as expected^2,4^. While some participants recorded no respiratory illnesses despite strong adherence, others recorded up to 16 distinct illness events. TempTracker also captured the previously described inter-individual variability in terms of illness duration (range 1-13 days) and symptom manifestation^2,4^, even when comparing responses to the same virus. Within COCOON, we identified 26 household transmission events with significant variability between family members in their symptom development/duration and virus positivity, as has been reported in other household transmission cohorts^38,39^. Deciphering the biological mechanisms underlying the observed variability between participants is one of the primary objectives of the AERIAL Study^5^, with our data supporting the use of a smartphone app to robustly characterise viral illness trends over time.

Across all collected swabs, viruses were detected in 67.9%, with rhinovirus the most prevalent, consistent with previous reports^40^. One limitation was our focus on eight common viruses, as additional viruses would likely be identified with a broader panel. However, our data indicates that the app effectively tracked variations in the seasonal prevalence of influenza A and RSV seen in the wider population (Figure 3). One important consideration is the impact that the COVID-19 pandemic had on altering viral transmission patterns within the Western Australian community during the AERIAL/COCOON study periods, such as the absence of detectable influenza A in 2021 as captured both in our study (Figure3A) and in public health data^41^. The pandemic restrictions also necessitated changes to the study protocol such as self-collection of swabs and reduced in-person participant contact. Overall, the ability of a smartphone app with self-collection of swabs to accurately characterise community viral transmission despite pandemic restrictions further highlight the strength of this approach, and its potential in other research and clinical contexts.

**Figure 2.**
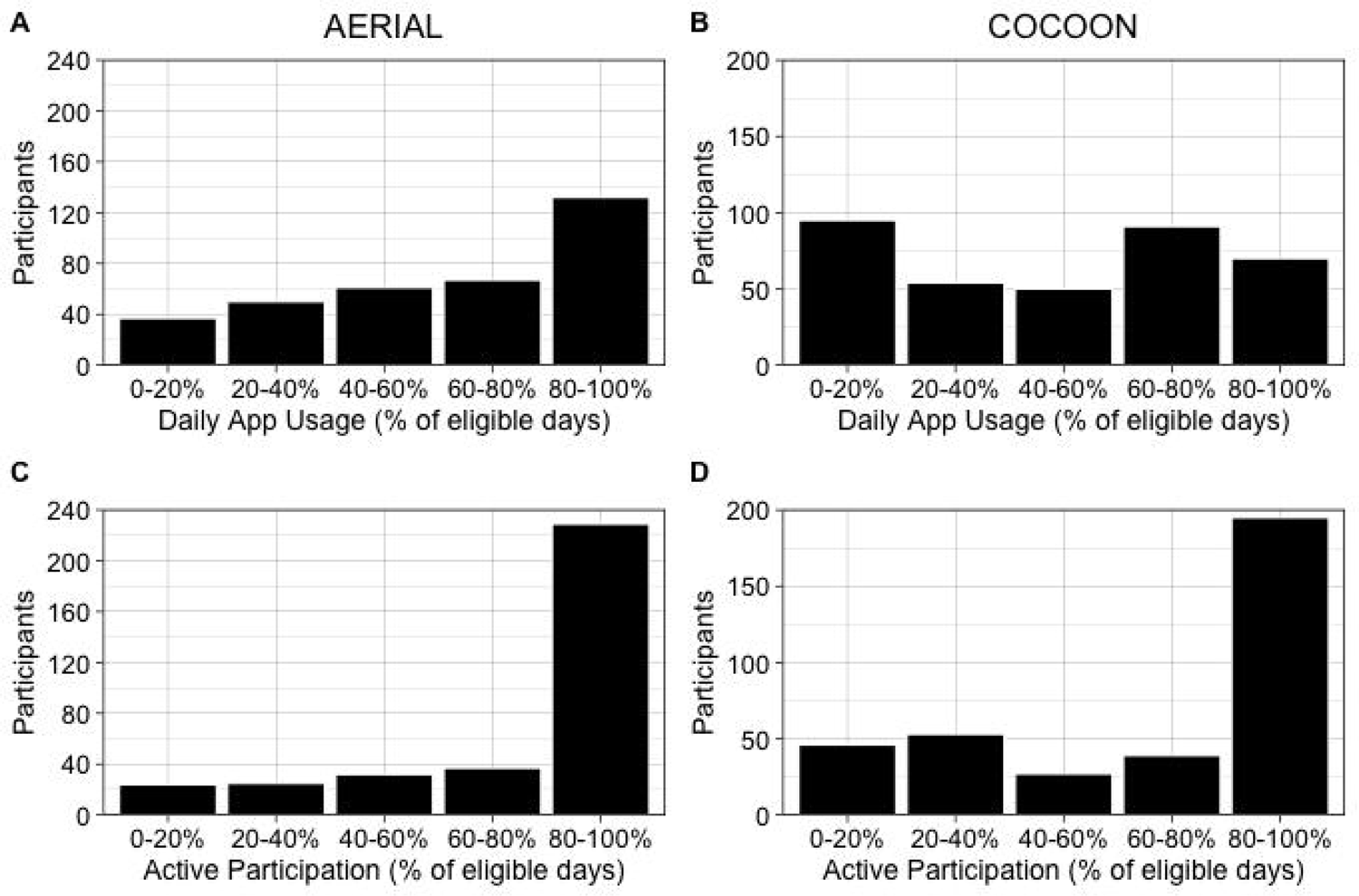
Adherence with AERIAL TempTracker app use in the AERIAL and COCOON cohort studies. Each participant’s eligible study period was calculated from the date they were initialised in the app through to 6-months or the date they were lost to follow-up/withdrew from the study. (A, B) Daily app usage was calculated as the percentage of days that the app was used out of the total number eligible study days for each participant in the (A) AERIAL and (B) COCOON studies. (C, D) “Active” participation was calculated as the percentage of days that a participant remained active in the app (i.e. using the app at least once every 7 days) out of the total number of eligible study days for each participant in the (C) AERIAL and (D) COCOON studies.

**Figure 3.**
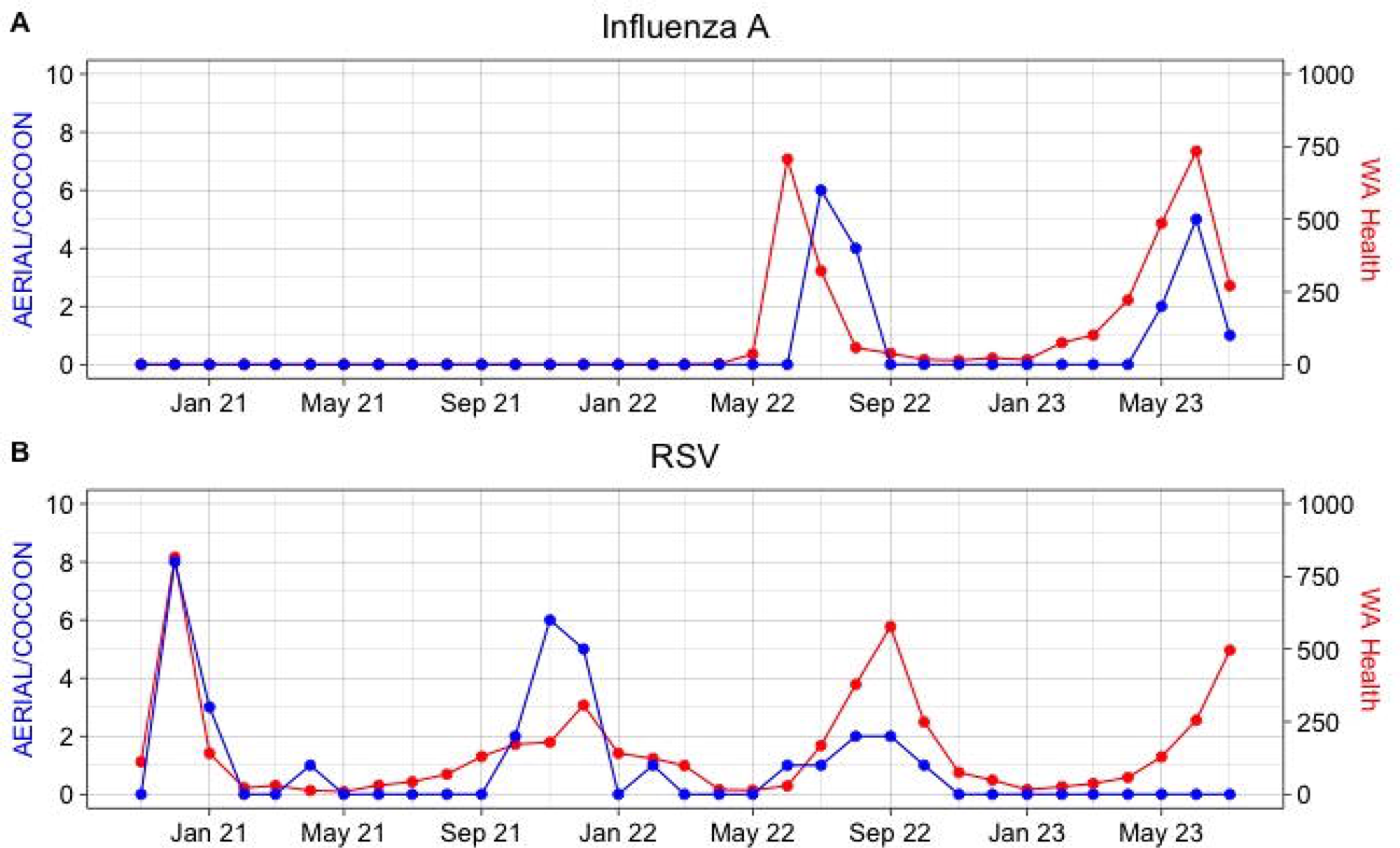
Influenza A and RSV prevalence in the AERIAL/COCOON studies matches virus transmission in the wider Western Australian community. The number of positive respiratory swabs for (A) Influenza A and (B) respiratory syncytial virus (RSV) from combined participants in AERIAL and COCOON were compared to Western Australian (WA) public health data from the same period.

While TempTracker was developed primarily to identify symptomatic respiratory illness, this smartphone-based approach could easily be adapted to other disease contexts. In support of this, most symptomatic events detected by TempTracker did not meet study criteria and instead captured other events such as gastrointestinal illness, allergy, or teething-related symptoms. Additional health information was also collected in our free text form, including potential vaccine reactions and other non-respiratory illnesses. Collectively these data support the extension and adaptation of comparable smartphone technology into other longitudinal cohort settings.

In summary, we have shown a smartphone app provides a reliable method to capture cohort-level respiratory virus data that mirrors community transmission patterns, with additional benefits including real-time adherence and symptom monitoring whilst reducing participant burden. We have also demonstrated the robust collection of longitudinal data of an individual participants’ illness burden, necessary to elucidate the mechanisms underlying phenotypic variability and vulnerability to respiratory viruses.

## Ethics statement

This study was performed in accordance with the Declaration of Helsinki. This human study was approved by Ramsay Health Care WA-SA HREC - approvals: #1908 & #2024. All parents, guardians or next of kin provided written informed consent for the minors to participate in this study. All adult participants provided written informed consent to participate in this study.

## Data Availability

All data produced in the present work are contained in the manuscript, however additional information /data are available upon request to the authors

## Acknowledgements

We would like to thank the contribution of the AERIAL and COCOON families, together with the cohort study investigators not listed as authors: Peter LeSouef, David Martino, Anthony Bosco, Susan Prescott, Desiree Silva and Asha Bowen. We also extend our gratitude to the hardworking and the dedicated research team, for the recruitment, liaising and sample collection over the duration of both cohort studies. The reported work was funded by NHMRC. SS holds an NHMRC Investigator Grant, TI is support edby the Stan Perron Foundation, AK is a Rothwell Family Fellow and DGH is a PCHF Fellow. Thes studies are a sub-project of The ORIGINS Project. This unique long-term study, a collaboration between Telethon Kids Institute and Joondalup Health Campus, is one of the most comprehensive studies of pregnant women and their families in Australia to date, recruiting 10,000 families over a decade from the Joondalup and Wanneroo communities of Western Australia. We are grateful to all the ORIGINS families who support the project. We would also like to acknowledge and thank the following teams and individuals who have made The ORIGINS Project possible: The ORIGINS Project team; Joondalup Health Campus (JHC); members of ORIGINS Community Reference and Participant Reference Groups; Research Interest Groups and the ORIGINS Scientific Committee; Telethon Kids Institute; City of Wanneroo; City of Joondalup; and Professor Fiona Stanley. The ORIGINS Project has received core funding support from the Telethon Perth Children’s Hospital Research Fund, Joondalup Health Campus, the Paul Ramsay Foundation, and the Commonwealth Government of Australia through the Channel 7 Telethon Trust. Substantial in-kind support has been provided by Telethon Kids Institute and Joondalup Health Campus.

