## Supplementary Data for "REAL TIME MONITORING OF RESPIRATORY VIRAL INFECTIONS IN COHORT STUDIES USING A SMARTPHONE APP"

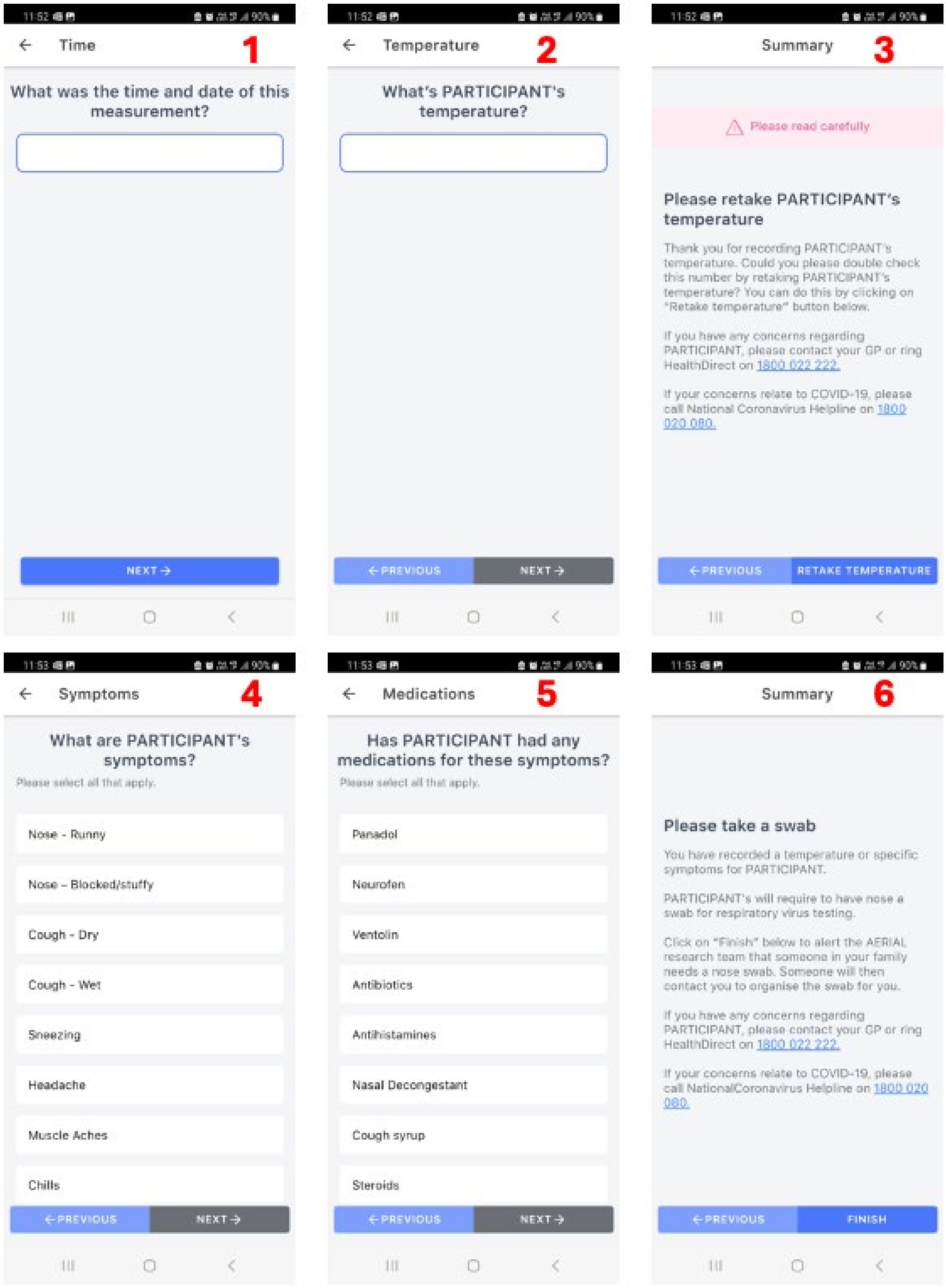


**Supplementary Figure 1.** TempTracker App. Participant's (or the parents of participants) entered the following data into the app: (1) the time/date and (2) the value of daily temperature measurements. (3) Participants were prompted to recheck and record temperatures >37.5. Participants were then asked to record (4) symptoms and (5) medications from a pre-defined list, with option to report other symptoms/medications in a free-text box. (6) Events meeting the case definition for a symptomatic respiratory illness generated an automated "Please take a swab" alert to participants and study staff.

**Supplementary Table 1:** Selected responses from consumers during testing of prototype.

| Supportive feedback | Feedback suggesting improvement |
|---|---|
| I deliberately put a high temp- the prompts to swab and instructions worked really well. | When you fill out a high temp and it asks you to enter it again it would be easier if you didn't have to refill the symptoms again. Not sure if that is possible. just seems a bit onerous when you have a sick child. |
| Looks smart but also simple and easy to use | I don't think it should be as easy to add a temperature from a past date. When looking at the screen if you are in a previous date it looks no different from todays view and it is easy to add a temperature reading without realising you are on the wrong day. |
| Great aerial app, easy to use, good clear explanations. | Perhaps a second reminder to take temp that day. If you are busy when the first reminder comes up you can't snooze it and no further reminders are given. |
| Was able to put all ID on many devices without a problem | When phone is on powersave mode, it does not prompt to take children's temp. I was only notified at 10pm when I plugged in the phone to charge before bed. |
| If no temps or symptoms have been recorded a reminder prompt is sent out | Download not the easiest – but I have a new phone so this probably didn't help. |

**Supplementary Table 2:** List of all symptoms and medications included in the TempTracker app. Respiratory symptoms for case definition denoted with *.

| SYMPTOMS | MEDICATIONS |
|---|---|
| NONE | NONE |
| Nose – Runny* | Panadol |
| Nose – Blocked/stuffy* | Neurofen |
| Cough – Dry* | Ventolin |
| Cough – Wet* | Antibiotics |
| Sneezing* | Antihistamines |
| Headache* | Nasal Decongestant |
| Muscle Aches* | Cough syrup |
| Chills* | Steroids |
| Repeated shaking with chills* | Other -> Free Text Entry |
| Tiredness* | |
| Sore Throat* | |
| Having trouble breathing* | |
| New loss of taste* | |
| New loss of smells* | |
| Feeding – not wanting* | |
| Not eating solids* | |
| Diarrhoea | |
| Crying | |
| Sleeping more | |
| Feeds – wanting more | |
| Cling | |
| Dribbling | |
| Rubbing at gums | |
| Pulling at ears | |
| Vomiting | |
| Nappies – less wet | |
| Nappies – more wet | |
| Rash | |
| Flushed cheeks/face | |
| Wind/colic | |
| Other -> Free Text Entry | |
